# A Fractal kinetics SI model can explain the dynamics of COVID-19 epidemics

**DOI:** 10.1101/2020.04.11.20061366

**Authors:** Kosmas Kosmidis, Panos Macheras

## Abstract

The COVID-19 pandemic has already had a shocking impact on the lives of everybody on the planet. Here, we present a modification of the classical SI model, the Fractal Kinetics SI model which is in excellent agreement with the disease outbreak data available from the World Health Organization. The fractal kinetic approach that we propose here originates from chemical kinetics and has successfully been used in the past to describe reaction dynamics when imperfect mixing and segregation of the reactants is important and affects the dynamics of the reaction. The model introduces a novel epidemiological parameter, the “fractal” exponent *h* which is introduced in order to account for the self-organization of the societies against the pandemic through social distancing, lockdowns and flight restrictions.

## Introduction

As of March 30, 2020, coronavirus disease 2019 (COVID-19) has been confirmed in 782,213 people worldwide, leading to 37,579 deaths. Its mortality is apparently higher compared with a mortality rate of less than 1% from influenza (*1*). There is an urgent need to model the growth of COVID-19 worldwide. The classical epidemiological approach for the study of growth relies on the reproduction number and infection time, which leads to an exponential growth. However, the data accumulated so far indicate deviation of growth from this pattern. Various approaches to model COVID-19 epidemics have been published in the literature recently based on various hypotheses (*2, 3*) Our model follows the principles of fractal kinetics (*4*) which is suitable for chemical reactions under “topological constraints” e.g. insufficient mixing of the reactant species. In fact, there is a full analogy between the governments measures to ensure social distancing for the control of epidemics and the reactions taking place in low dimensions or insufficient stirring (*4–6*)

## The Model

The classical SI Model of epidemic spreading is the simplest approach in the mathematical modeling of epidemics (*7, 8*). A population comprises two classes, susceptible and infected. The fraction of susceptible individuals is denoted by *S* and that of the infected is denoted by *I*. The total number of individuals is assumed to be constant and consequentially the sum of the fractions is equal to one, i.e. *S* + *I* = 1. Thus, the SI model is essentially reduced to a single non linear ordinary differential equation

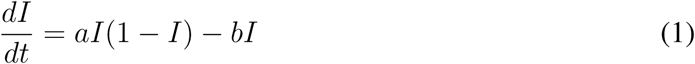

where *a* is proportional to the probability of an infected individual to infect a health one and *b* is the recovery rate of infected individuals. In the standard SI model it is common to write the *a* constant per person, i.e. *a/N* where *N* is the total size of the population but for our present investigation the form of eq 1 is adequate.

There are several drawbacks in this model. Most importantly the parameter *a* is considered a constant, while as recent experience has shown in global epidemics societies tend to organize, governments take measures to ensure social distancing etc.

The fractal kinetic approach that we propose here originates from chemical kinetics and has successfully been used in the past to describe reaction dynamics when imperfect mixing and segregation of the reactants is important and affects the dynamics of the reaction (*4, 9, 10*)

A fractal kinetics approach consists in assuming that the reaction constants are in fact functions of time. In particular the assumed form is obtained if *a* → *a/t*^*h*^. The power law form of the function and the “fractal” exponend *h* is the reason of the term “Fractal Kinetics” proposed by Kopelman in his famous article (*4*).

Thus, the fractal kinetics SI model for epidemic spreading is described by the following equation

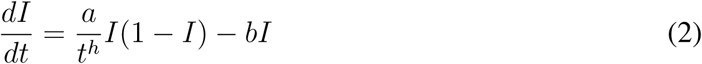

Although it is in principle possible to fit the above equation to the available data for the spreading of the COVID-19 epidemic and calculate the three parameters *a, b, h*, in practice this approach is rather difficult since equation 2 can only be solved numerically and numerical instabilities when *t* is close to zero cause problems to the fitting process. A better approach is to consider the cumulative fraction of infected individuals *I*_*T*_ as a function of time. The differential equation describing this quantity according to the Fractal Kinetic SI model would be similar to equation 2 with the omission of the recovery term, thus

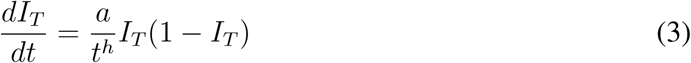

This equation cal be solved analytically and one obtains the rather elegant result

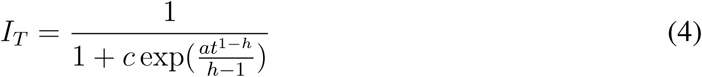

This equation can be fitted to the confirmed cases for each country as a function of time and as we present below the fitting is excellent in all cases. Here, we focus on the study of the exponent *h* which is the novel significant parameter introduced in the scope of Fractal kinetics. High values of the exponent *h* signify high level of self-organization of the system, while the value of *h* = 0 reduces the model to the classical SI model where perfect mixing of the population is assumed. The parameter *c* is of considerable importance as it determines the asymptotic limit of *I*_*T*_ i.e. the total fraction of individuals that will be infected. Taking the limit of eq 4 when *t* → ∞ we find

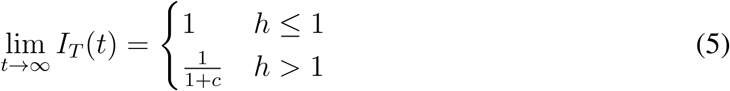

It is well known, however, that most countries test only for serious symptomatic cases and thus the number of confirmed cases seriously underestimates the actual fraction of infected people. The parameters *a* and *c* of the model are rather sensitive in such systematic underestimation while the parameter *h* is rather robust. In order to demonstrate that we have estimated the *a, c, h* parameters for the actual data for Greece and then we have multiplied the data by 10^3^ and recalculated them. We found that *c* changed by a factor of approximately 10^3^, the relative of *a* was almost 40% while the exponent *h* changed only by roughly 7% i.e from 1.37 to 1.27.

## Results

Figure 1 shows results for the confirmed cases of nine countries. In each case the confirmed cases are normalized by dividing them by the country population. For each country day zero is considered to be the day of the first confirmed case. Points are the COVID-19 data and the solid line is the model’s best fit of eq 4 to the data (*11*).

**Figure 1:**
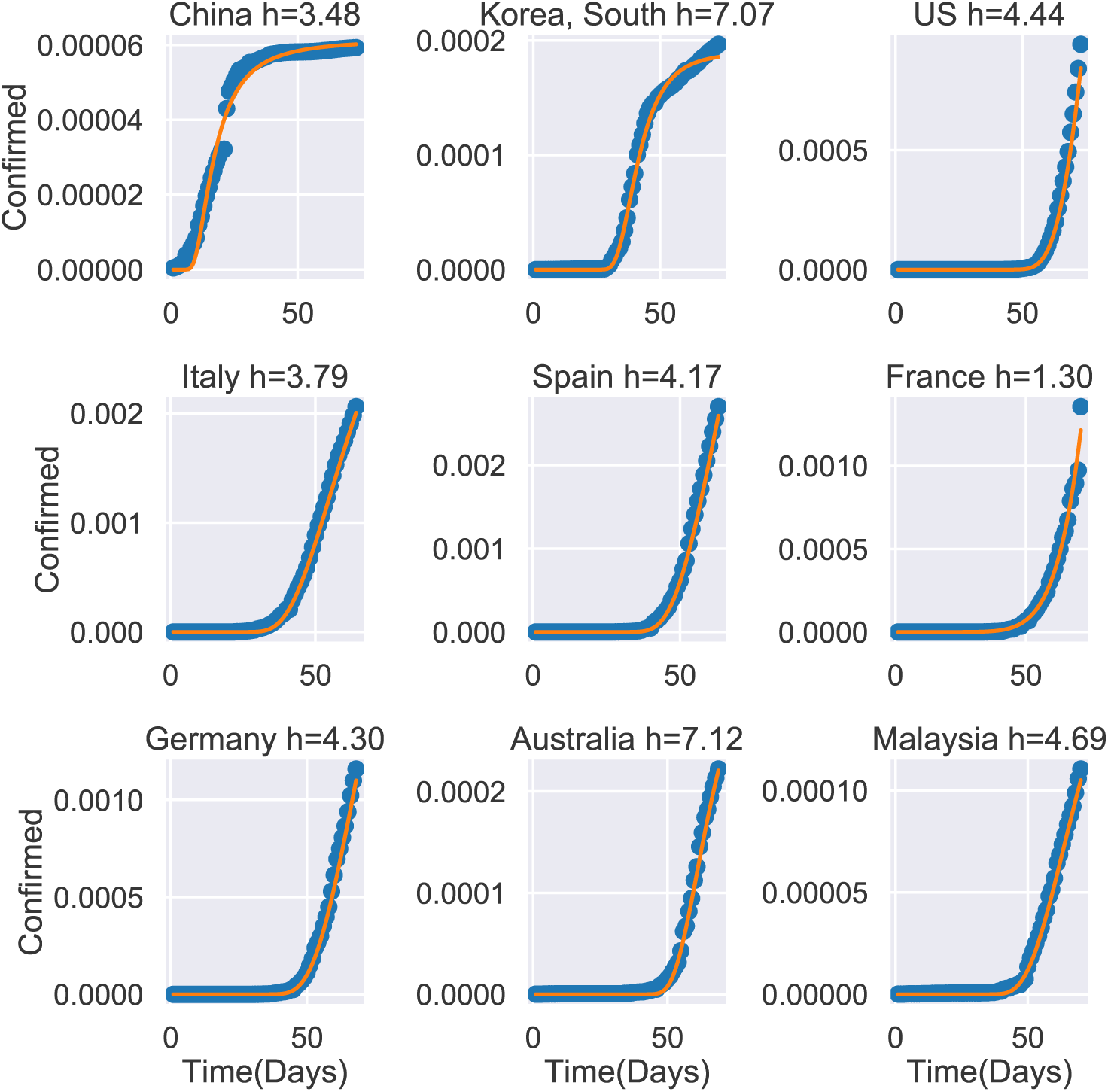
Confirmed cases of COVID-19, *I*_*T*_ as a function of time for 9 countries. Points are the actual data (*11*). The solid line is the best fit of of eq 4.Data up to 4-4-2020.

Figure 2 shows countries with more that 1000 confirmed cases ranked in a decreasing order of the *h* exponent as calculated using outbreak data up to the 29 March 2020. Australia and South Korea are ranked on top with practically the same *h* exponent. For South Korea this is in agreement with the recognition this country has received from the international press on the way that has reacted to the COVID-19 epidemic outbreak (*12*). For Australia the high *h* value is probably a result of the country’s preventive measures.

**Figure 2:**
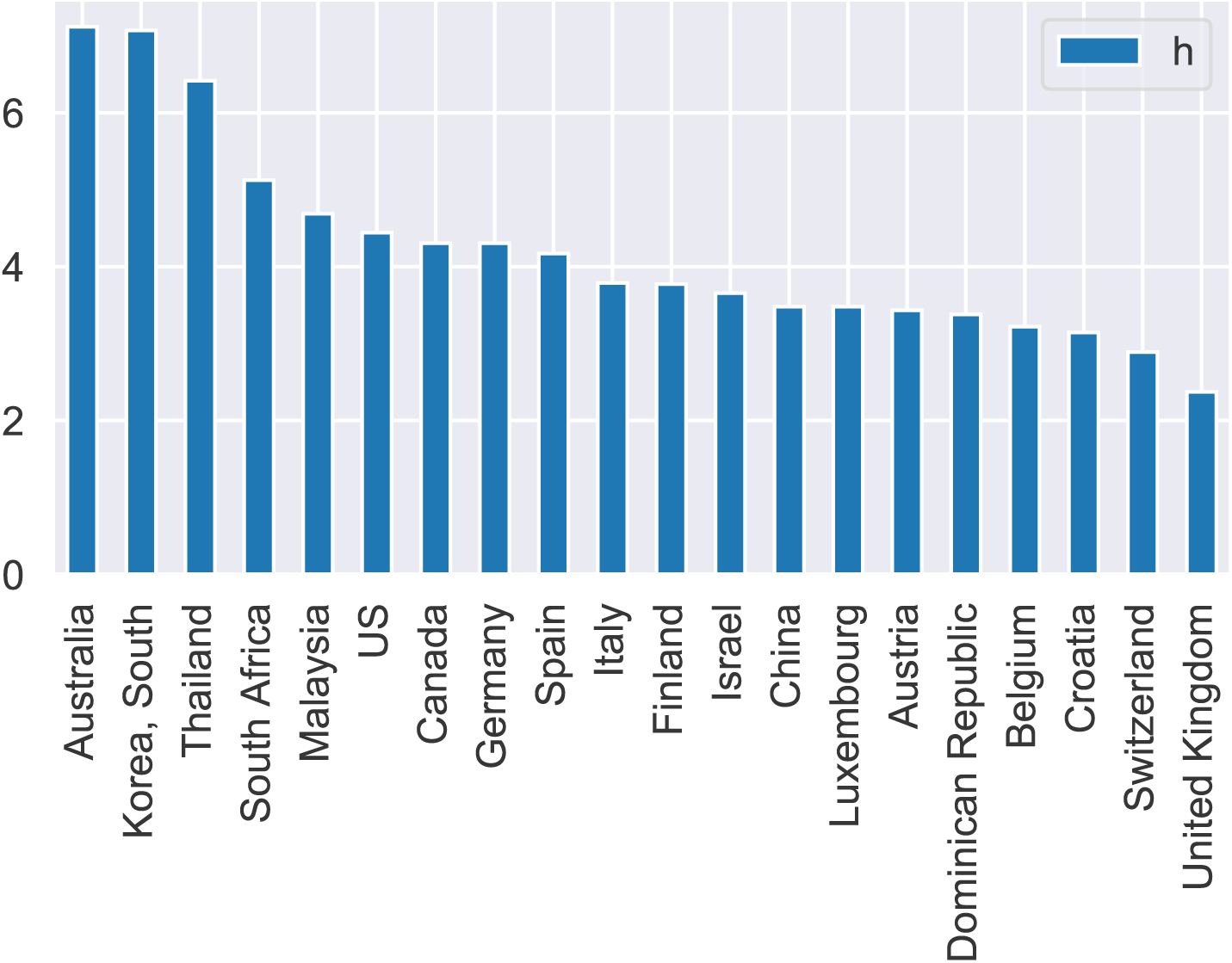
Ranking of Countries (top 20) with more that 1000 Cases according to the calculated value of the *h* exponent. Data up to 4-4-2020.

We expect that the calculated values of the *h* exponent will change as new data become available and as measures of increasing “social distancing” become effective. Due to the long time of the virus incubation period it is believed that “social distancing” measures taken today will have an observable effect in the number of confirmed cases in 2 weeks. Interestingly, using data up to 29-3-2020 the estimated *h* exponent for Italy is equal to 2.76 while using the latest data (up to 4-4-2020) the Italian *h* exponent increases to 3.79 after the dramatic increase in outbreak cases in Italy and the extreme prevention measures taken by the Italian government (*13*).

Here we proposed a simple model based on fractal kinetics that is in excellent agreement with the published COVID-19 outbreak data. Of course more detailed modeling could reveal more aspects of the outbreak and lead to a better understanding. We feel, however, that the exponent *h* of the fractal kinetic SI model is a novel, and easily determined parameter, from an epidemiological point of view and, thus, can provide new insight to the disease dynamics. Moreover, due to the excellent fit of Eq. 4 to the data for all countries, we believe that even more detailed models would ultimately lead to an expression for the fraction of infected individuals *I*_*T*_ that is similar (if not identical) to Eq. 4.

Finally, it would be helpful to identify commonalities in the set of parameters *a, c, h*. Thus, we assume that each country is epidemiologicaly characterized by the three component vector *a, c, h*. Since Eq. 4 is an approximation it is anticipated that these three parameters are correlated. Thus, we use an unsupervised learning method, namely a Principle Components Analysis (PCA), a statistical procedure that uses an orthogonal transformation to convert a set of observations of possibly correlated variables into a set of values of linearly uncorrelated variables called principal components (*14*). Figure 3 shows a plot of the two largest PCA components.

**Figure 3:**
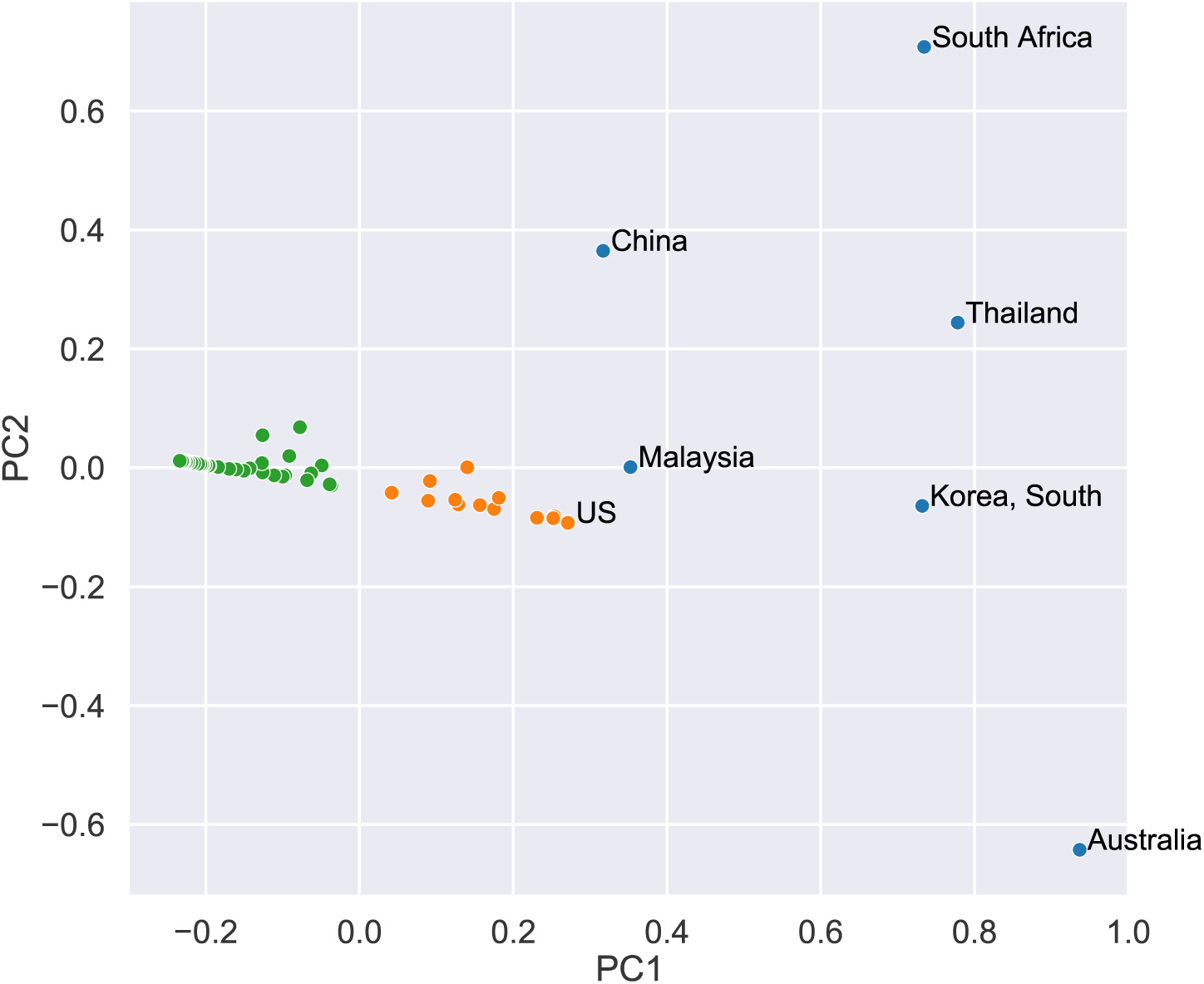
Plot of the largest *PC*1 vs the second largest *PC*2 principle components for countries with more than 1000 confirmed cases. Each country is characterized by its *a, c, h* vector of the Fractal SI model. The principle components were calculated with data up to 4-4-2020. For clarity only the names of countries with *PC*1 *>* 0.26 are shown.

We observe roughly 3 clusters-colored by green, orange and blue. The majority of countries have a negative value of *PC*1 and a small value of *PC*2. There is second cluster of countries with positive *PC*1 and small *PC*2 value, including countries like US, Canada and Germany.

The 3rd cluster comprises countries with high values of *PC*1 like South Korea, Malaysia and Australia which are also countries with large *h* exponent. The major goal of this work is to propose efficient ways to model the epidemic spreading when taking under account the “heterogeneity” that arises as a response of the societies to the epidemic. The fractal kinetic framework, is a simple, almost naive extension of the classical models which seems to work surprisingly well.

## Data Availability

All data in the manuscript are publicly available data

